# Real-time genomics to track COVID-19 post-elimination border incursions in Aotearoa New Zealand

**DOI:** 10.1101/2021.05.13.21257194

**Authors:** Jordan Douglas, Jemma L. Geoghegan, James Hadfield, Remco Bouckaert, Matthew Storey, Xiaoyun Ren, Joep de Ligt, Nigel French, David Welch

## Abstract

There have been thirteen known COVID-19 community outbreaks in Aotearoa New Zealand since the virus was first eliminated in May 2020, two of which led to stay-at-home orders being issued by health officials. These outbreaks originated at the border; via isolating returnees, airline workers, and cargo vessels. With a public health system informed by real-time viral genomic sequencing which typically had complete genomes within 12 hours after a community-based positive COVID-19 test, every outbreak was well-contained with a total of 225 community cases, resulting in three deaths. Real-time genomics were essential for establishing links between cases when epidemiological data could not, and for identifying when concurrent outbreaks had different origins. By reconstructing the viral transmission history from genomic sequences, here we recount all thirteen community outbreaks and demonstrate how genomics played a vital role in containing them.

**Summary:** The authors recount the role of real-time viral genomics in containing the COVID-19 community outbreaks of Aotearoa New Zealand.

## Introduction

From early on in the COVID-19 pandemic, New Zealand (Aotearoa in te reo Māori) has adopted an elimination approach to the disease. Elimination was first achieved in May 2020 (Cousins, 2020; Baker et al., 2020) and from then to 30 April 2021 only thirteen community outbreaks (**Table 1**) comprising a total of 225 community cases have been recorded. We define a community case to be someone who has either been in contact with the wider community while potentially infectious, or was infected after being put into a managed isolation and quarantine (MIQ) facility due to one of the outbreaks discussed here. Community cases are distinct from MIQ cases who are returnees who only came into contact with others in MIQ during their infectious period.

**Table 1:**
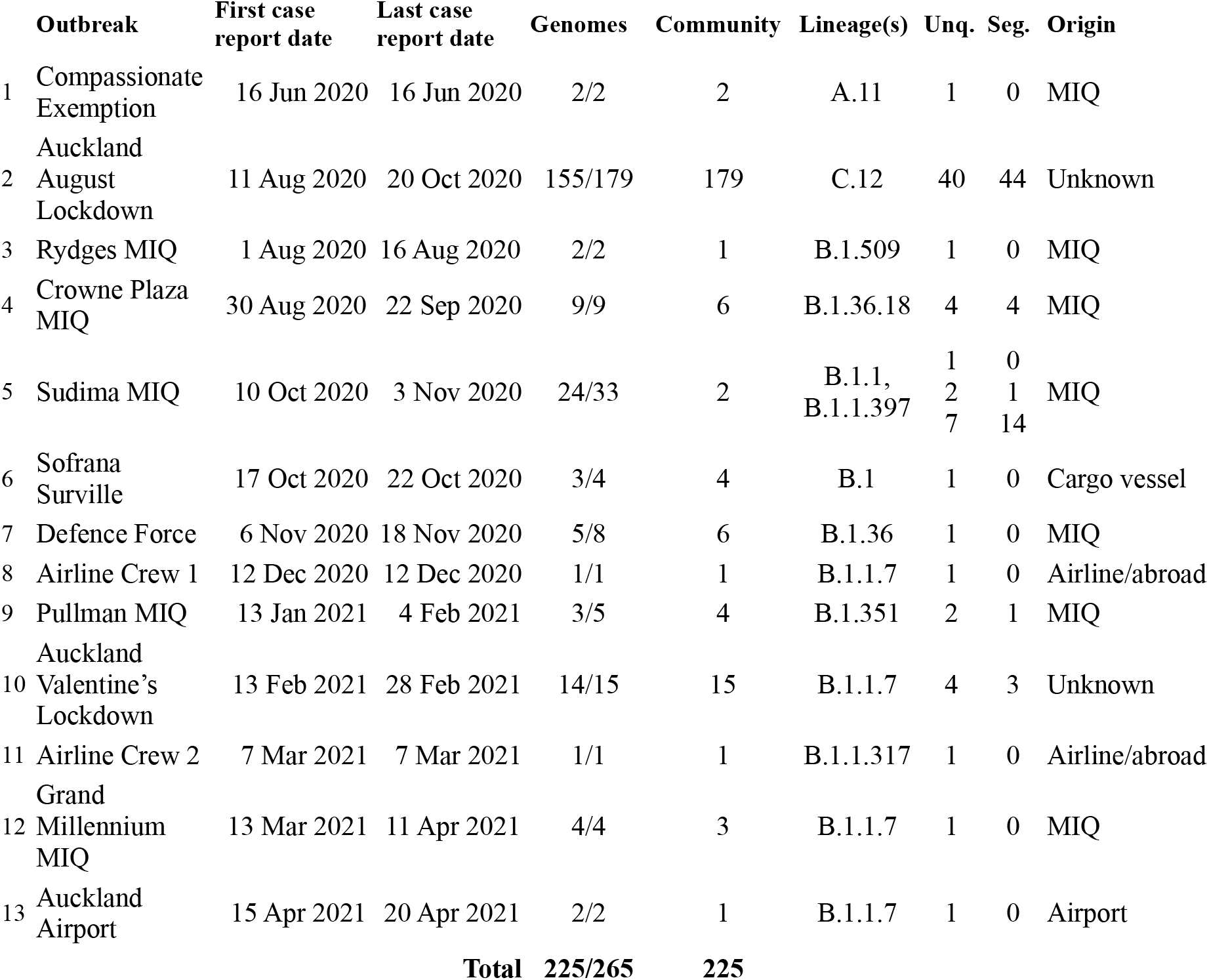
Summary of all COVID-19 community cases in New Zealand since elimination was first achieved. Pangolin lineages are specified (version 2.3.9; Rambaut et al. (2020)) as well as the number of complete genomes / number of confirmed cases, and the number of community cases. The number of unique genomes (Unq.) and the number of segregating sites (Seg. i.e. the number of genome positions which differ) within each cluster are counted, or within the three sub-clusters of the Sudima outbreak. All dates are the dates of report for the lab test.

The public health response to community outbreaks have differed depending on their extent. Two outbreaks resulted in Auckland, Aotearoa’s largest city, moving to Alert Level 3 that mandates stay-at-home-orders for most people under the Alert Level system (comprising Levels 1 to 4, with 4 being the most stringent; Jefferies et al. (2020)).

A core part of the elimination strategy is a strictly controlled border where nearly every person entering the country is required to isolate for 14 days at an MIQ facility, and be tested for SARS-CoV-2 at days 0, 3, and 12 of their stay (Jefferies et al., 2020; New Zealand Government, 2020). The MIQ facilities are repurposed hotels, over half of which are located in Auckland. With an operational capacity of 4000 returnees, the returnees (135,451 to date; MBIE, 2021) and the considerable workforce required to service them present a possible transmission route into the community. Of the thirteen known border incursions, seven originated in MIQ facilities (four from MIQ workers, and three from returnees who tested positive after leaving the facility), three were airline workers, one was from an infection on a visiting ship, while the sources of the remaining two, which both led to stay-at-home orders, are still unknown.

Since 19 April 2021 there has been an open border between Aotearoa and Australia. Australia has pursued a similar elimination strategy and uses a hotel based MIQ system, though with notably fewer but larger outbreaks detected from their MIQ facilities (Grout et al., 2021; Smith, 2020).

Viral genomic sequencing has played a crucial role in tracing and understanding all community outbreaks in Aotearoa (Geoghegan et al., 2020, 2021c), complementing border controls, the Alert Level system, and contact tracing. There has been an effort to sequence the virus from every case. Routine cases or infected returnees in MIQ are sequenced weekly. Community or otherwise suspect cases are sequenced more urgently, with complete genomes typically being available to inform health officials within 12 hours of the first positive test. This real-time genomic surveillance has been indispensable in confirming or disproving links between cases, particularly when epidemiological data was lacking.

In the remainder of this article, we recount the events surrounding the thirteen community outbreaks to date, and demonstrate how genomic sequencing technologies have played vital roles in delimiting them.

## Outbreaks

We reconstructed the phylogenetic tree for Aotearoa’s border incursions, using complete viral genomes from Aotearoa and, for context, from the rest of the world (Shu and McCauley, 2017). The subtrees showing the border incursions are presented in **Fig. 1**, while the full global tree can be found in **Supporting Information.**

**Fig. 1:**
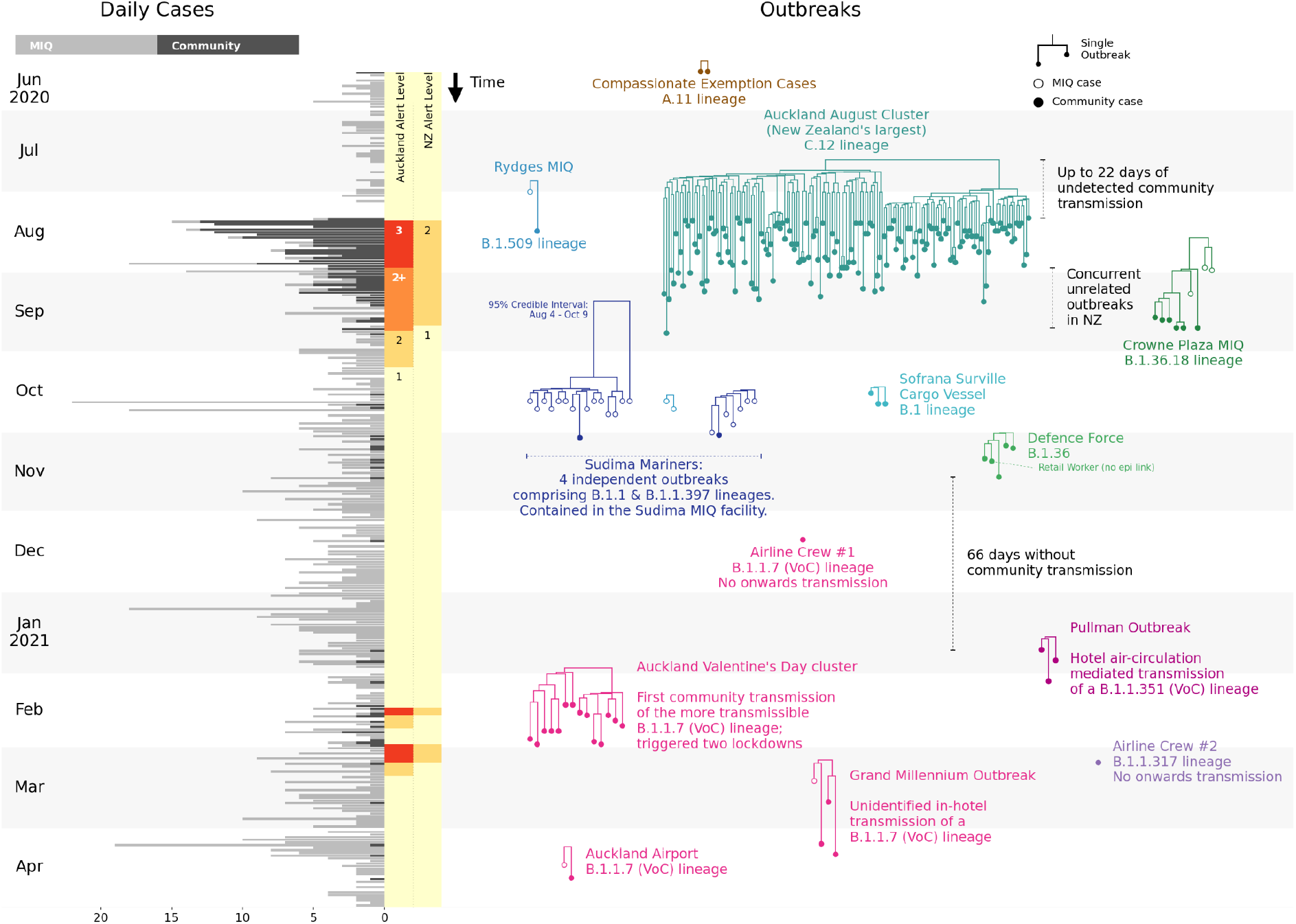
Left: daily COVID-19 cases in Aotearoa New Zealand, with the Alert Levels in the Auckland region and across the wider country indicated. Right: phylogenetic trees of all thirteen COVID-19 post-elimination community outbreaks, with variants of concern (VoC) indicated. Each subtree displayed here is part of the larger phylogenetic tree built from genomes around the world.

As of 30 April 2021, complete genomes (>90% recovery) have been obtained from 1288 out of 2243 cases in total (57%), and 583 of 1030 cases since 1 June 2020 (57%). Some cases without a full genome sequence had lower quality genomes available which were sufficient to assign them to a lineage, but many had insufficient viral material for any meaningful analysis. For the community outbreaks considered here, we have high quality genomes for 85% (225 of 265) of the cases (**Table 1**).

### Compassionate Exemption

After 24 days without any recorded cases in the community or at the border, and one week after the whole of Aotearoa had moved down to Alert Level 1, two cases were found in the community. They had arrived on 7 June and were granted a compassionate exemption to exit MIQ early in order to attend a funeral on 13 June. The conditions of the exemption required them to self-isolate as far as possible while travelling and to get tested. They both tested positive for COVID-19 on June 16. Within one day, complete viral genome sequencing confirmed that the two cases shared a single origin. Although there were no secondary infections, concern for public health saw the New Zealand Defence Force put in charge of managing MIQ facilities, no returnees being allowed to leave MIQ without first returning a negative test, and the end of compassionate exemptions from most of the MIQ requirements.

### Auckland August Lockdown

On 11 August 2020, four cases of COVID-19 were found among workers at an Auckland cold storage facility ending a 102 day period with no recorded community transmission. The city was sent into an immediate lockdown (Alert Level 3), and lower level restrictions were introduced for the rest of the country (Alert Level 2). Elevated restrictions remained until 7 October as the cluster grew to a total of 179 cases, including 3 deaths. This was the largest COVID-19 cluster in New Zealand.

All of the genomes were closely related with 44 segregating sites found across 155 genomes (**Table 1**). The single origin provided confidence to public health officials that it was a single outbreak despite several cases being found that had no clear epidemiological links with other cases (NZMH, 2020b). The cluster included a healthcare worker who was infected during their work at an MIQ facility where community cases were sent to quarantine (NZMH, 2020e). While the index cases worked at a cold chain supply facility linked to the border, the source of the outbreak was never established (Geoghegan et al., 2021a).

Complete genomes are available for 87% of cases in this outbreak which we believe makes it one of the most comprehensively sampled large COVID-19 outbreaks.

### Rydges MIQ Facility

During the Auckland August outbreak, there was an unusual case of a maintenance worker at the Rydges MIQ facility with no known epidemiological link to the ongoing community outbreak. Sequencing confirmed that the source of infection was not related to the main community cluster, but rather to an overseas returnee under managed isolation at the Rydges MIQ facility. A followup investigation suggested that transmission likely occurred when the two cases used the same elevator in close succession (NZMH, 2020f) and was likely a case of airborne transmission (van Rijn et al., 2020; Dbouk and Drikakis, 2021).

### Crowne Plaza MIQ Facility

The complete Crowne Plaza outbreak has been thoroughly analysed in Eichler et al. (2021) so we provide only brief detail here. In September 2020, a returnee developed symptoms and tested positive in Auckland four days after leaving the Crowne Plaza MIQ facility in Christchurch. Genomic sequencing showed that this case (“Case G”) – and their two household contacts – were not linked to the ongoing Auckland August cluster. Rather, they were linked to other returnees under managed isolation at the Crowne Plaza. Epidemiological investigations show the chain likely started with Cases A and B who were seated close to Case C on a repatriation flight from India. C went on to infect D in the MIQ facility via airborne transmission between a hotel room and its adjacent hallway. D is then thought to have infected G on a domestic flight from Christchurch to Auckland. This outbreak illustrates the power of genome sequencing – when coupled with detailed epidemiological investigations – for identifying cryptic transmission events such as those on flights or airborne transmission between MIQ guests. The genomic evidence here is complete with all sequences in the cluster available and the tree relatively well-resolved with four distinct genomes among its nine cases.

### Sudima MIQ Facility

235 international mariners arrived into New Zealand on a charter flight from Moscow on 16 October 2020 and began their self-isolation at the Sudima MIQ facility in Christchurch. In the ensuing period, 31 of the mariners tested positive for COVID-19 as did two workers at the MIQ facility (NZMH, 2020g). Genomic sequencing produced full genomes for 24 cases and indicated at least four independent origins behind the 33 cases, falling into three distinct phylogenetic clades (**Fig. 1**). The three clades consist of two, seven, and 15 cases. We estimate the origin of the largest clade being between 4 August and 9 October (95% credible interval), but the mariners arrived on 16 October, suggesting there were at least two separate introductions of this variant into the facility. The two MIQ workers were infected with two different variants of the virus, with public health officials concluding that the two transmission events occurred via regular interactions with the mariners where protocols were followed but were insufficient to prevent transmission (Venter Consulting, 2020).

### Sofrana Surville

In mid October, a border worker tested positive for COVID-19 as part of routine testing. After extensive surveillance testing, three others tested positive; two were household contacts and one was a mariner who worked onboard the same cargo vessel – the Sofrana Surville (NZMH, 2020h). Genomic testing confirmed that the four cases shared the same origin, and were part of a lineage that was novel to New Zealand, thus making residual community transmission an unlikely explanation. International crew members onboard the Sofrana Surville were confirmed as the source of infection when the ship arrived in Australia and crew were tested by Australian health officials in Queensland. One crew member tested positive and the sequence was reported to match the New Zealand cases.

### Defence Force

After visiting a number of public locations in the Auckland central business district, a New Zealand Defence Force member tested positive. Genomic sequencing confirmed his infection to be acquired from the quarantine facility he worked at. Contact tracing identified three additional cases in Wellington, and surveillance testing identified one community case in the Auckland central business district. The only known connection to the rest of the outbreak was that this case worked at a retail outlet about 50 metres from one of the locations the index case visited (NZMH, 2020c). The case was quickly linked to the rest of the cluster through whole genome sequencing, providing reassurance that widespread undetected community transmission was unlikely (NZMH, 2020d). The circumstances of the transmission event remain unknown. Because the genomic link was established, there was no change in Alert Level. However, the general public were asked to avoid the Auckland central business district where possible for about three days.

### Airline Crew 1

An airline crew member tested positive for COVID-19 after a flight from the United States of America. They were detected within 48 hours and there were no recorded secondary infections (NZMH, 2020a). Genomic sequencing indicated they acquired their infection abroad.

### Pullman MIQ Facility

A returnee tested positive for COVID-19 one week after completing managed isolation at the Pullman MIQ facility. Despite extensive travel across the Northland region whilst infected with the B.1.351 lineage (Tegally et al., 2020; Cele et al., 2021), there were no secondary infections reported, including their travelling companion. Shortly thereafter, two additional cases with the same variant we found in the community, both of whom had completed self-isolation at Pullman (NZMH, 2021c). The outbreak was successfully limited to these four community cases who were genomically linked to a returnee under isolation at Pullman. The epidemiological link was thought to be the air circulation system at the MIQ facility. New arrivals at the MIQ facility were suspended and its air filtration systems were improved.

### Auckland Valentine’s Lockdowns

On the eve of Valentine’s day 2021, three household members in Auckland tested positive. The genomes were identical and of the highly transmissible B.1.1.7 variant (Volz et al., 2021; Davies et al., 2021). Although one case worked at an airline services company, they had no obvious contact with people coming through the border. Auckland was immediately sent into its third Alert Level 3 lockdown for a period of three days, and wide scale surveillance testing commenced. Other cases with closely related genomes were found, however the epidemiological links were not always strong (NZMH, 2021d). A fourth Alert Level 3 lockdown followed shortly afterwards when a further case was found that could not immediately be linked to the cluster.

The known outbreak was restricted to four households and 15 cases, although epidemiological gaps suggest there may have been undetected cases. With genetic evidence showing four distinct genomes among 14 cases, the genomics were not informative beyond confirming a single origin, and the B.1.1.7 variant’s global overrepresentation presented further difficulties at pinpointing an overseas origin. The outbreak has not been traced back beyond the original three cases and its origin remains unknown.

### Airline Crew 2

During the Auckland Valentine’s outbreak, an airline crew member tested positive a week after their arrival from Japan (NZMH, 2021a). Their infection was likely acquired overseas and there were no known secondary infections.

### Grand Millennium MIQ Facility

In late March 2021, a cleaner at the Grand Millennium MIQ facility tested positive. Despite being infected with the more transmissible B.1.1.7 lineage, there were no known secondary infections in the community. Two weeks later, two further workers from the same facility also tested positive (NZMH, 2021b). All three cases were genomically linked back to a returnee isolating at the Grand Millennium.

### Auckland Airport

A worker at Auckland International Airport tested positive for the B.1.1.7 variant in April 2020. No onward transmission was detected. Genomic sequencing linked the case to a recent returnee, and a followup investigation suggested that worker’s infection was obtained from cleaning the airplane the returnee had arrived on, and was likely a case of airborne transmission (NZMH, 2021e).

## Discussion

Genomic sequencing has been used in real-time to investigate each of Aotearoa New Zealand’s thirteen post-elimination community outbreaks. Sequencing has been essential not only for establishing links between cases when epidemiological links could not (e.g. the Defence Force outbreak, and both Auckland lockdowns), but also in identifying when multiple outbreaks had different origins (e.g. decoupling the Rydges outbreak from the ongoing Auckland outbreak, and the second Airline crew case from the Valentine’s outbreak). These efforts have been instrumental in clearly delineating outbreaks and informing the public health response. Genomics have also elucidated cryptic modes of transmission such as airborne transmission (Eichler et al., 2021) and in-flight transmission (Swadi et al., 2021), which have brought about policy changes (e.g. revision of filtration systems in MIQs).

When paired with routine genomic sequencing from within MIQ facilities and around the world, genomics can identify the origins of community outbreaks and rule out the possibility of undetected widespread community transmission. However, as exemplified by the two community outbreaks that led to lockdowns, this strategy has limited ability to identify outbreak origins when there is no closely matched genome with a plausible epidemiological link (Geoghegan et al., 2021a).

A wide range of lineages have been imported into Aotearoa over the course of the pandemic (**Table 1**, Geoghegan et al., 2020). The fact that the four locally acquired outbreaks in 2021 were all caused by variants of concern (VoC) – Pangolin lineages B.1.1.7 and B.1.351 (Wang et al., 2021a,b; Rambaut et al., 2020) – reflects the lineages that are arriving at the border, with 83 out of the 142 genomes from overseas returnees found from 1 January to 30 April 2021 being from those two lineages or other VoCs. However, there is too little data here to make any link between these outbreaks and the reported higher transmissibility of the VoCs.

New Zealand’s ability to rapidly generate SARS-CoV-2 genomes has greatly improved over the past year, to the point where new genomes are routinely available within hours of positive community tests. Although the potential of the techniques described here have been well characterised in academia (Gardy and Loman, 2018), the pandemic has facilitated widespread adoption in New Zealand and other places (such as Singapore and Australia (Pung et al., 2020; Seemann et al., 2020)); with the term “whole genome sequencing” becoming commonplace in public health announcements. Combined with epidemiological investigation, these data have increased public understanding of the outbreak and driven policy change. Although the techniques described here of real-time sequencing and analysis coupled with epidemiological investigation have come to the fore during the COVID-19 pandemic, they are not limited to pandemic situations. These technologies can be integrated into regular surveillance of other pathogens, such as seasonal influenza viruses, which have been largely absent from many countries over the past year (Olsen et al., 2020).

## Supporting information

GISAID acknowledgements

## Data Availability

Sequence data available on GISAID

## Materials and Methods

We constructed a multiple sequence alignment (Katoh and Standley, 2013) containing 225 genomes from New Zealand community outbreaks and a further 663 from the rest of the world, downloaded from GISAID (Shu and McCauley, 2017). For each New Zealand outbreak we sampled up to 50 global sequences from the same Pangolin lineage(s) (Rambaut et al., 2020) as the outbreak, uniformly through time between the date of the first case in the outbreak and 60 days before it. In order to reduce the effect of geographical sampling biases, global sequences were down weighted proportional to the number of sequences from the same country. For example, in order to sample around the Pullman MIQ outbreak, we considered all B.1.351 global genomes collected up to 60 days before the outbreak, and sampled 50 genomes from this pool, where the probability of sampling genome x was inversely proportional to the number of genomes in the pool from the same country as x. Our tree is the maximum clade credibility tree summarising a posterior distribution of trees inferred by BEAST 2 (Bouckaert et al., 2019). Genomic sites were partitioned into the three codon positions, plus non-coding, as described by Douglas et al. (2020). For each partition we modelled evolution with an HKY substitution model with log-normal(μ=1, σ=1.25) prior on kappa, frequencies estimated with Dirichlet(1,1,1,1) prior, and relative substitution rates with Dirichlet(1,1,1,1). We use a strict clock model with log-normal(μ=-7, σ=1.25) prior on mean clock rate, and for the tree prior we use a Bayesian skyline model (Drummond et al., 2005) with Markov chain distribution on population sizes and log-normal(μ=0, σ=2) on the first population size. We established convergence of the analysis by running multiple (8) analyses and using Tracer (Rambaut et al., 2018) to ensure effective sample sizes were sufficient and all individual analyses converged to the same distribution. See the XML file in **Supporting Information** for further details.

