## Supplementary figures and images for "Real-time genomics to track COVID-19 post-elimination border incursions in Aotearoa New Zealand"

### Supplemntal figure 1

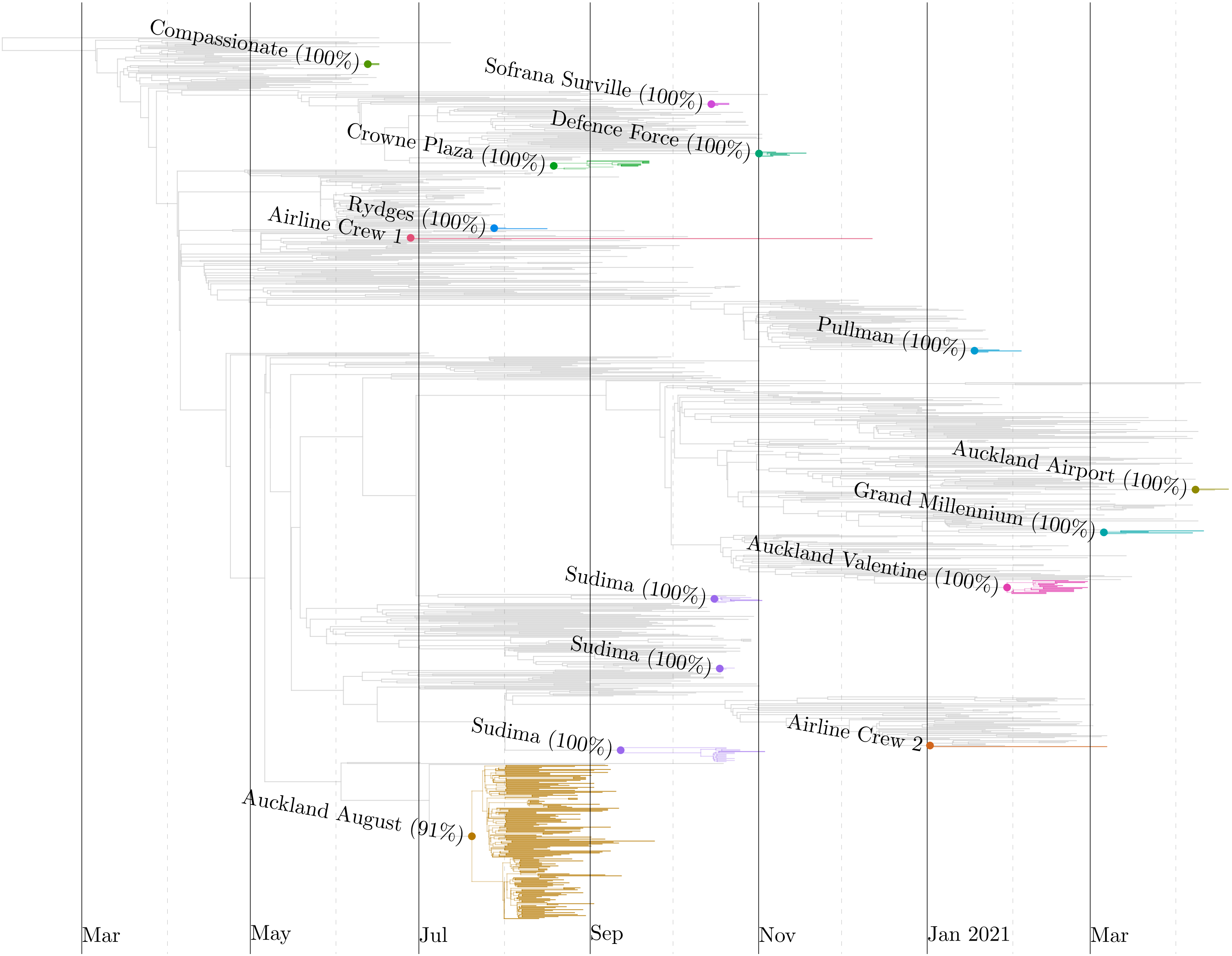
